# Gene regulatory network analysis identifies dysregulation of hypoxia pathways as contributing to glioblastoma multiforme treatment resistance in females

**DOI:** 10.64898/2026.01.13.26344041

**Authors:** Tomisin Adebari, Viola Fanfani, Marouen Ben Guebila, Derrick DeConti, Katherine Hoff Shutta, Camila M. Lopes-Ramos, Lauren Hsu, Dawn L. DeMeo, John Quackenbush, Tara Eicher

## Abstract

**Background:** Glioblastoma multiforme (GBM) is an aggressive brain tumor that is notoriously resistant to treatment, with an average survival time of 17 months. While the overall outcome is poor for both males and females, sex differences in GBM incidence and outcome suggest sex-specific biological mechanisms underlie tumorigenesis. In contrast, low-grade glioma (LGG) is a less aggressive brain tumor that tends to have a better prognosis and a longer survival time.

**Methods:** To understand mechanisms contributing to treatment resistance in GBM in both males and females, we inferred gene regulatory networks (GRNs) for males and females with LGG and GBM using RNA-seq data from The Cancer Genome Atlas (TCGA). We analyzed these to identify both sex-specific and sex-stratified gene regulation in GBM.

**Results:** We found few sex-specific differences in gene regulation in individuals with LGG, consistent with the lack of evidence for significant clinical endpoints dependent on sex. However, in GBM-we found sex-specific differential targeting of several pathways, including hypoxia and related pathways (carbohydrate metabolism, innate immune processes, and extracellular matrix pathways) known to be dysregulated in hypoxic conditions. In comparing between individuals with GBM, we found that females exhibited a greater degree of co-regulation between hypoxia with the aforementioned downstream pathways than did males.

**Conclusions:** Our results suggest that dysregulation of hypoxia-related pathways in GBM plays a female-specific role in resistance to treatment and overall outcomes.

## Background

Gliomas originate in the glial cells that support neurons and contribute to roughly 26% of all brain tumors and 80% of malignant brain tumors (1). The World Health Organization (WHO) classifies gliomas based on pathological evaluation, with grades 1 and 2 considered low-grade gliomas (LGG) and grade 4 corresponding to glioblastoma multiforme (GBM) (2). GBM is associated with poor prognosis and is resistant to both chemotherapy and radiation therapy (3), having a median survival of about 17 months with multimodality therapy and 3 months without any form of treatment (4). LGG has a far better prognosis than GBM, with a median survival of about 7 years (5).

Treatment outcomes are poor for both males and females with GBM; although females respond better to treatment with temozolomide (the standard of care) than males (6-8), the five-year GBM survival rate among females is still less than 10% (6). This difference in treatment response motivates the question of whether, and if so how, GBM treatment resistance mechanisms differ between the biological sexes. In contrast to the sex-bias in GBM, numerous studies have found that an individual’s sex is not associated with outcome in LGG, despite a slightly higher incidence in males (9, 10). Although sex-specific molecular features have been reported in GBMs, such as *MGMT* hypermethylation in females (11), other factors contributing to sex differences in GBM treatment resistance remain poorly understood (12, 13). This suggests that jointly analyzing GBMs and LGGs may help shed light on the mechanisms driving the clinically relevant sex differences observed in GBM.

A common approach to differential analysis in molecular biology is to compare DNA sequence and gene expression levels between groups (14), sometimes in combination with other data such as methylation, but this may miss vital biological correlations or regulatory relationships (15). Gene regulatory network (GRN) inference can provide valuable context regarding the regulatory relationships that define expression patterns and the characteristics of phenotypic states (15). Although people often use the term GRN to refer to a wide range of possible network associations, we define GRNs as the inferred associations between regulators and their targets—most often transcription factors (TFs) and the genes they regulate. GRN inference and analysis has proven particularly useful in the study of sex differences in health and disease. For example, in analyzing gene expression in postmortem brain tissue from nominally healthy individuals in the Genotype-Tissue Expression (GTEx) Project (16), we no meaningful differences in gene expression between males and females. However, when we used Passing Attributes between Networks for Data Assimilation (PANDA) (17) to infer GRNs, we found that up to 36% of genes were targeted by different TFs in males and females (18). In other contexts, GRNs inferred using PANDA have \ provided deep insight into the drivers of biological systems (15, 19, 20), including identifying PD1 signaling as a predictor of poor prognosis in GBM (21).

We used PANDA to infer GRNs using autosomal and allosomal gene expression from The Cancer Genome Atlas (TCGA) (22), with separate networks for each of the following four groups: (1) GBM samples from females (GBM-F), (2) GBM samples from males (GBM-M), (3) LGG samples from females (LGG-F), and (4) LGG samples from males (LGG-M). PANDA infers regulatory networks between TFs and target genes by using message passing to find consistency between a TF-gene prior based on mapping transcription factor binding sites to regulatory regions across the genome, likely TF-TF complexes based on protein-protein interaction data, and data on pairwise gene co-expression that reflects likely co-regulation of genes (23, 24). The resulting GRN is a bipartite graph that links TFs and their target genes via “edges” that reflect the strength of evidence for a regulatory TF-gene relationship. In the PANDA GRNs, genes that are associated with particular states are those that exhibit substantial changes in their “targeting score,” which we define as the sum of all edge weights corresponding to candidate regulatory TFs. We performed this analysis comparing GBM-F with GBM-M, LGG-M with GBM-M, and LGG-F with LGG-M. For each, we used the ranked list of differentially targeted genes and performed gene set enrichment analysis to identify functional gene sets and pathways within these groups. We also used Modeling Network State Transition from Expression and Regulatory data (MONSTER) (25) to identify differences in GBM-specific (relative to LGG) TF activity by sex, comparing LGG-F with GBM-F and LGG-M to GBM-M. Finally, we used Bipartite Limited Subnetworks from Multiple Observations using Breadth-First Search with Constrained Hops (BLOBFISH) (26) to find subnetworks connecting differentially targeting TFs within each PANDA network. See Figure 1 for an overview.

**Figure 1.**
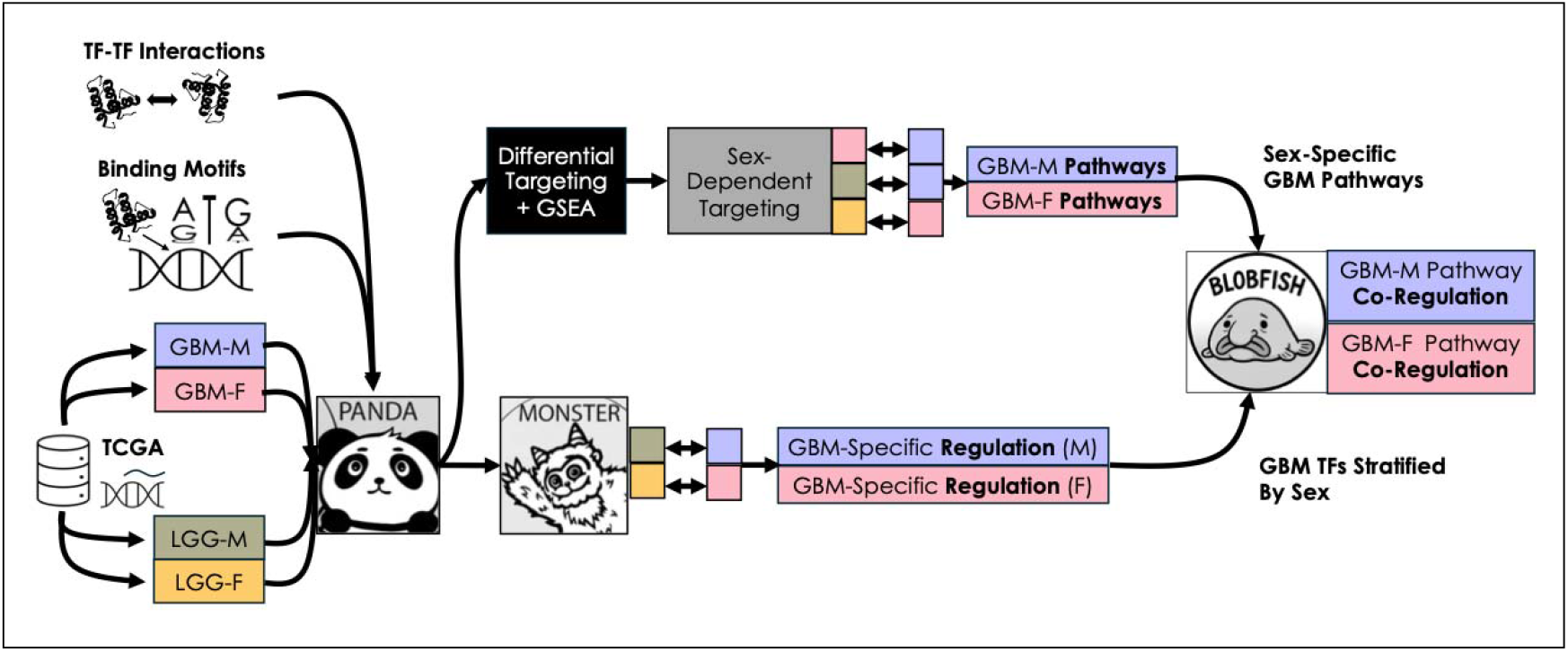
Analytical workflow. Double arrows indicate comparisons between subgroups.

Using this integrated strategy, we identified TFs, genes, and pathways that differed between the sexes and were specific to later-stage GBM—the disease state that exhibits the most significant phenotypic difference between the sexes. Among the sex-specific pathways we found in GBM are several related to hypoxia, mRNA splicing, the extracellular matrix, immune processes, and carbohydrate metabolism. Further, we found that TFs contributing significantly to GBM in males or females differentially co-regulate these pathways, indicating sex-specific mechanisms that regulate essential disease processes. Although previous studies have explored sex-specific differences in GBM (8, 27) using gene expression (28), methylation (29), and mutation (30), our findings provide unique insight into the regulatory mechanisms driving the differences between GBM and LGG in each sex, and provide a list of candidate targets for sex-specific treatment in GBM.

## Methods

We inferred GRNs from TCGA data stratified by sex and tumor type and identified GBM-specific regulation in males and females, which we then used to characterize the biological mechanisms underlying GBM-specific treatment resistance in males and females. Scripts used in this analysis are available at https://github.com/QuackenbushLab/Adebari_Glioma_scripts.

### Data preprocessing

We obtained TCGA GBM (*n* = 153) and LGG (*n* = 652) RNA-seq raw count data for autosomes and allosomes, and clinical annotation data from WebMeV, an open-source web-based application that uses cloud computing resources to provide users with access to genomic analysis tools and large public-domain datasets (31). We normalized raw counts using *voom*() (Variance Modeling at the Observational level) in the R Bioconductor package *limma* (32), which transforms RNA-seq count data into log2-counts per million (logCPM). We filtered the normalized data to include only protein-coding genes as annotated in the Genome ENCyclopedia (GRCh38.p14) (33) to simplify the dataset, reduce noise, and facilitate interpretation. We stratified the normalized and filtered data by tumor type and sex: LGG-F (n = 273), LGG-M (n = 379), GBM-F (n = 53), and GBM-M (n = 100).

### Gene regulatory network inference

We used the Python version of PANDA (24) (available from the netZooPy repository v0.10.3, https://github.com/netZoo/netZooPy) (34) to estimate aggregate GRNs for each stratum. The resulting GRN is a bipartite graph connecting TFs to their target genes, with edge weights indicating the confidence that a TF regulates a gene. We used sex-specific priors generated as described by Saha *et al*. (35) but where, in females, prior weights for TFs specific to the Y chromosome were set to 0. The motifs were derived from the Catalog of Inferred Sequence Binding Preferences (CIS-BP) database (36), built on the human genome build GrCh38. We downloaded PPI from the Gene Regulatory Network Database (GRAND) (37). Using PANDA with these prior and PPI networks, we constructed GRNs for LGG-F, LGG-M, GBM-F, and GBM-M co-expression (Supplementary Tables S1-S4).

### Differential targeting and pathway analysis

To examine regulatory mechanisms specific to GBM in males and females, we identified sex-specific differences in gene targeting between the LGG and GBM strata and between GBM-M and GBM-F, but not between LGG-M and LGG-F. We chose to compare GBM to LGG rather than to a healthy control to better understand treatment-resistance mechanisms specific to GBM, given that LGG has a better response and does not exhibit strong differences between the sexes. We calculated a gene targeting score,

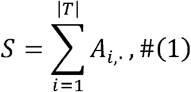

where *S* is the vector of targeting scores for all genes within a stratum, *A* is the weighted adjacency matrix of the GRN, with rows corresponding to TFs and columns corresponding to target genes, and *T* is the set of all TFs represented in the GRM.

The gene targeting score for each gene in each GRN represents the extent to which that gene is targeted by TFs in that GRN. Because PANDA GRNs are not guaranteed to include the same range of edge weights, we ranked the gene targeting scores in each stratum for cross-strata comparison. In male-female comparisons (GBM-M to GBM-F and LGG-M to LGG-F), we removed GRN edges in which targets were Y chromosome genes.

For each of these comparisons, we computed the difference in rankings (differential targeting) between the two conditions as defined in Equation 2, where *Rank(S)* is the ranking of *S, S*_*1*_ corresponds to condition 1 (for example, GBM-F), *S*_*2*_ corresponds to condition 2 (for example, GBM-M), and *D* corresponds to the difference in rankings.

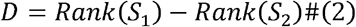

Using the *fgsea* R package (38) a significance threshold of FDR-adjusted *p*-value < 0.05 to identify pathways enriched for differentially targeted genes, with pathway annotation from the Human Molecular Signatures Database (v2023.2) canonical pathways collection (which includes pathways curated by BioCarta, Kyoto Encyclopedia of Genes and Genomes (39), Pathway Interaction Database (40), Reactome (41), Sigma-Aldrich, UCSD Signaling Gateway, and WikiPathways (42)).

From these, we identified GBM-specific pathways differentially targeted in females, defined by pathways enriched in both the GBM-F to GBM-M and GBM-F to LGG-F comparisons, but not the LGG-F to LGG-M comparison. Similarly, we identified GBM-specific pathways differentially targeted in males, defined by pathways enriched in both the GBM-M to GBM-F and GBM-M to LGG-M comparisons, but not in the LGG-M to LGG-F comparison. As noted previously, we used LGG as a comparison to identify regulatory mechanisms specific to GBM that are absent in LGG.

### Transcription factor activity between LGG and GBM in males and females

We used MONSTER (25) (available from the netZooR repository https://github.com/netZoo/netZooR) to identify key TFs associated with differences between GRNs generated in different contexts. MONSTER is based on a model of cell state transitions between a first GRM represented as a *p×m* adjacency matrix *A* (for *p* genes and *m* TFs) into a second, related GRN represented by as a *p*×*m* adjacency matrix *B* through as,

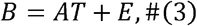

where *T* is an *m×m* transition matrix that “rewires” the network in ways that change connectivity, activates or inactivates different processes, and subsequently alters phenotype, and *E* is a *p×m* error matrix. MONSTER uses a regression-based method to estimate the transition matrix *T* from which one can deduce the regulatory drivers of biological state transitions, defined as those in *T* with the greatest off-diagonal weight. We used MONSTER to identify TFs rewired between LGG and GBM in both males and females, comparing GRNs between LGG-F and GBM-F, and between LGG-M and GBM-M. MONSTER scored each TF based on its overall contribution to the GRN differences between LGG and GBM in each sex. We manually annotated these TFs with functions; UniProtKB/Swiss-Prot (43) summary field information as captured in GeneCards (44).

### Co-regulation of sex-specific pathways by GBM-specific transcription factors

We wanted to explore the connections between the GBM-specific, sex-biased TFs and genes in the context of the GRN model networks (LGG-M, LGG-F, GBM-M, and GBM-F) described above, to identify subnetworks linking the TFs and to find cooperative effects of TFs in regulating key pathways. We used Bipartite Limited Subnetworks from Multiple Observations using Breadth-First Search with Constrained Hops (BLOBFISH) (26) (available from the netZooR repository). BLOBFISH models each node in a bipartite graph as having a local *sphere of influence*, defined by a breadth-first search limited to a user-specified number of hops. Two nodes are considered connected if they share nodes within these spheres, allowing multiple connecting paths that reflect redundancy and shared function, as seen in biological networks. To ensure robustness, BLOBFISH retains only subnetworks in which all edges have statistically significant weights relative to a null model. Because the graph is bipartite, all relevant connecting paths can be identified by examining the shared nodes equidistant from each pair of seed nodes.

We filtered each of the four PANDA networks to include only TFs with significantly different activity between LGG and GBM in either males or females, as determined by our MONSTER analysis. As target genes, we selected all GBM-specific pathway genes that were differentially targeted in either males or females based on our differential targeting analysis. We set the maximum hop length in BLOBFISH to 1 to capture only TFs that had evidence of co-regulating the input genes while excluding other genes. We considered the edges specific to GBM-M and GBM-F as evidence of coordinated regulation in each sex. The goal of this analysis was to uncover sex-stratified co-regulatory processes which, when considered in tandem with sex-specific regulatory processes and regulatory processes shared between sexes, may be used to inform the development of therapeutics.

## Results

### Hypoxia-related GBM-specific pathway targeting differs between males and females

The differential targeting analysis in the upper branch of Figure 1 allowed us to identify GBM-specific and sex-specific REACTOME and other pathways, as summarized in Figure 2; differentially targeted pathways are color-coded to reflect the sex in which we observed enrichment, and a manually assigned classification of the pathway category is provided.

**Figure 2.**
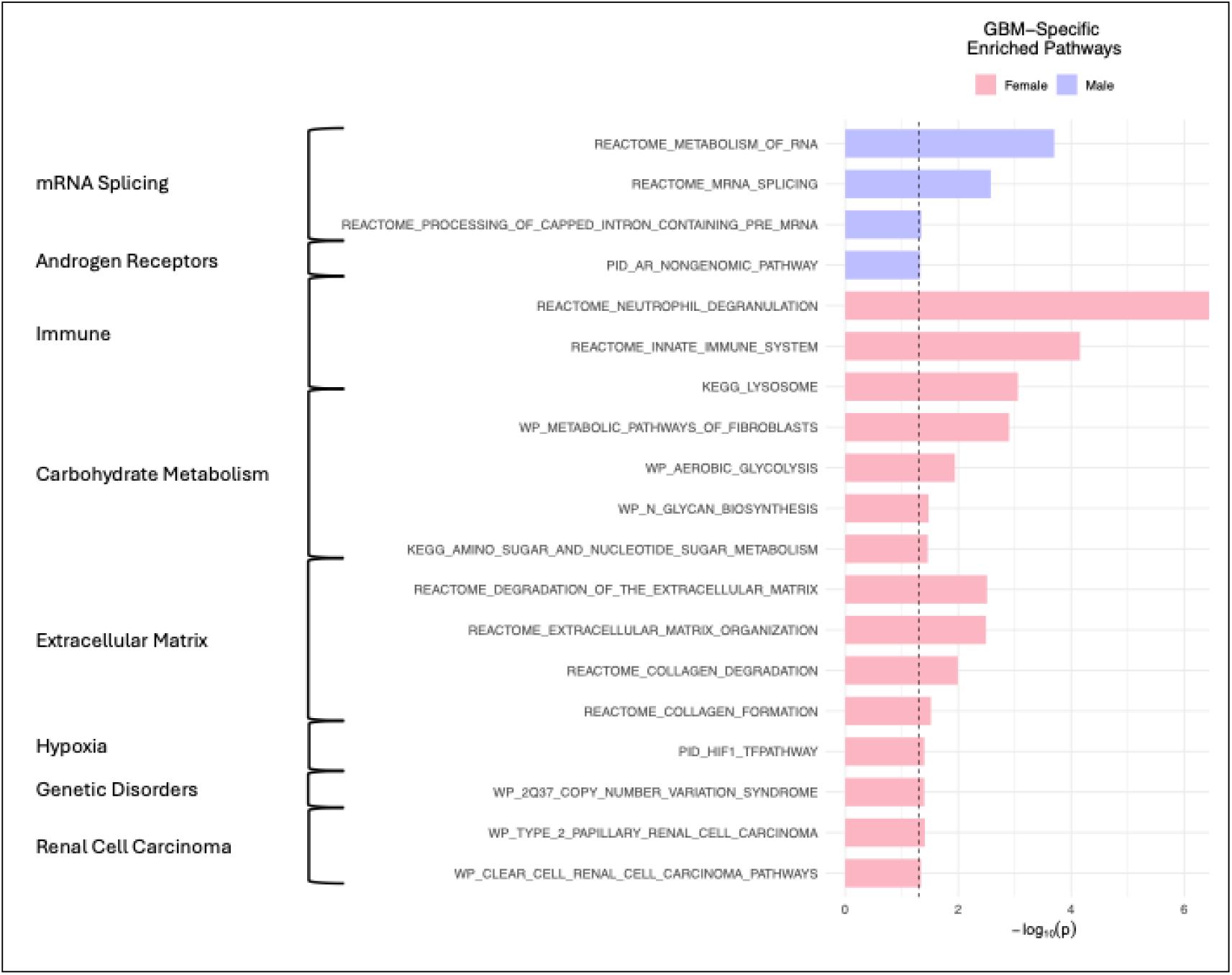
Enriched MSigDB pathways that are GBM-specific and sex-specific for males or females. Pathway enrichment was computed using *fgsea* with an FDR-adjusted *p*-value cutoff of 0.05.

**Figure 3.**
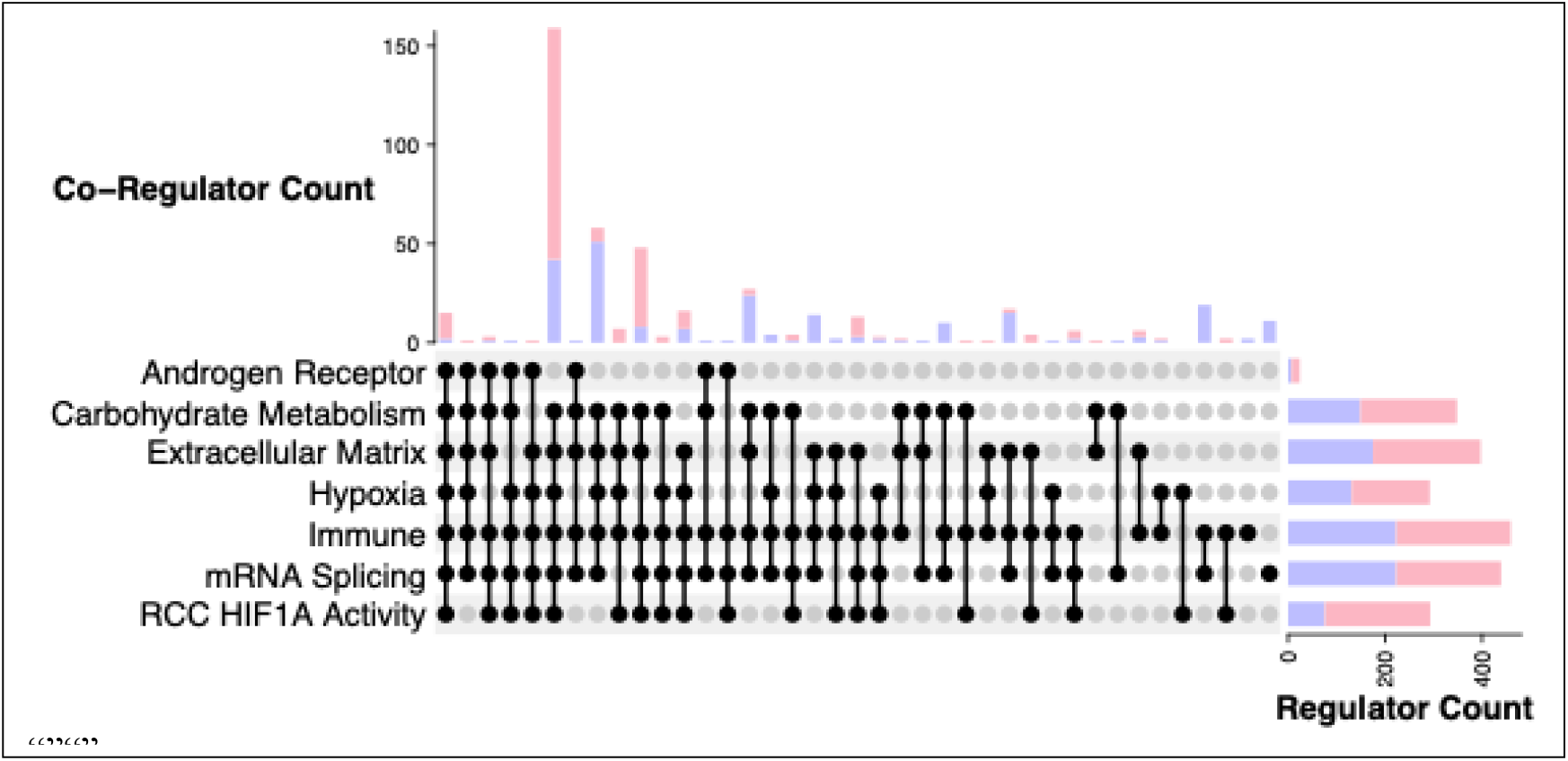
Sex-specific co-regulation obtained using BLOBFISH. Co-regulation of differentially targeted pathways in female (red) and male (blue) GBMs. “Regulator Count” refers to the number of TFs regulating unique genes in each pathway category, and “Co-Regulator Count” refers to the number of TFs co-regulating unique genes in multiple pathway categories. The filled circles in each column represent the sets of co-regulated pathway categories.

Pathways differentially targeted in GBM in females include immune response pathways, carbohydrate metabolism, extracellular matrix formation, and pathways associated with renal cell carcinoma (RCC). In males, we identified distinct pathways in GBM, including the spliceosome and androgen receptor pathways.

While RCC may at first glance appear to be spurious, the GSEA score was driven by differential targeting of genes involved in several processes, including angiogenesis (*VEGFA* and *PDGFRA*), glycolysis (*LDHA, PGK1, PSAT1, GAPDH, SHMT2, TPI1, SDSL, PSPH, ENO2, HK3, PKM, GPI, ALDOA, LDHB*, and *LDHC*), cell proliferation (*CAMK1* and *PLOD2*), cell migration and invasion (*AKT1S1, RHEB*, and *MLST8*), and the tumor suppressor *VHL*, which is involved in the ubiquitination of *HIF1A*, itself a key factor in hypoxia known to activate glycolysis, angiogenesis, and cell proliferation in RCC (45). This result is consistent with the other female-specific pathway targeting. The activation of glycolysis is represented by differential targeting of aerobic glycolysis and glycan biosynthesis in the broader “Carbohydrate Metabolism” category. Likewise, angiogenesis is represented by differential targeting of fibroblast, extracellular matrix, and collagen pathways in the broader “Extracellular Matrix” category (46, 47). Moreover, the differential targeting of neutrophil degranulation and lysosomal pathways within the broader “Immune” category is consistent with the known role of hypoxia in tumor-associated lysosomal dysfunction (48), all of which suggest a key role of hypoxia in glycolysis, immune evasion, and angiogenesis in GBM in females. In contrast to the GBM-specific dysregulation of hypoxia and downstream pathways in females, differentially targeted GBM-specific pathways in males included mRNA splicing and an androgen receptor pathway, with genes involved in the spliceosomal E, pre-B, B, B^act^, B*, C*, and P complexes, all of which affect the enrichment scores, and lead to the conclusion that different mechanisms are at play in GBM treatment resistance in males. A more complete pathway enrichment analysis is presented in Supplementary Tables S5-S12.

### Females exhibit differential targeting of *HIF1A* and other regulators of hypoxia response in GBM in comparison to LGG

Using MONSTER, we identified 249 TFs that differentially targeted genes between LGG and GBM in females and 12 in males (Full MONSTER results are presented in Supplementary Tables S13-S14). Of these, 240 were found only in females only and three only in males (zinc finger proteins *ZNF225, ZNF287, ZNF570*). Among those found only in females were 12 TFs known or predicted to regulate innate immunity or immune response, 10 associated with immune cell differentiation, and three with response to hypoxia (*CXXC5, EGFR*, and *HIF1A*) (Supplementary Table S15). This result further supports a female-specific role of hypoxia regulation in GBM, either in combination with or in tandem with the dysregulation of immune processes.

### Hypoxia is co-regulated with carbohydrate metabolism, extracellular matrix, and immune pathways in females only

We wanted to build on the differential targeting analysis to identify co-regulatory mechanisms that contribute to treatment resistance in GBM in males and females, so we reduced the PANDA GRNs to include only the TFs that differentially target genes between GBM and LGG, in either males or females, as well as their targets. We defined the extent of co-regulation as the number of TFs targeting genes in each pathway category and searched for co-regulators of multiple sex-specific pathway categories.

We found that the female-specific regulatory subnetwork (Supplementary Table S16) had evidence of more extensive co-regulation between hypoxia pathways and carbohydrate metabolism, the extracellular matrix, and immune pathways than did the male-specific regulatory subnetwork. This result suggests that the downstream activation of these pathways by *HIF1A*, a GBM-specific TF in females but not in males, is facilitated by dysregulation of both hypoxia and the downstream pathways that are under regulatory control by the same TFs. In contrast, the male-specific regulatory subnetwork (Supplementary Table S17) included co-regulation between mRNA splicing and the carbohydrate metabolism, extracellular matrix, and immune pathways, but not co-regulation of hypoxia. This provides strong evidence that GBM differs between males and females through patterns of TF regulation and co-regulation of critical biological processes.

## Discussion

The difference in regulation of genes involved in hypoxia between the sexes and its importance in GBM but not LGG may help to explain the difference in GBM treatment response and outcomes between the sexes. Hypoxia is known to increase resistance of tumors to chemotherapy (49). This occurs in part due to metabolic changes that occur in cancer cells in the presence of hypoxia (50), including carbohydrate metabolism pathways. In GBM specifically, *HIF1A* promotes angiogenesis, glucose metabolism, and migration (51). Moreover, hypoxia can contribute to immune evasion via multiple mechanisms (52). Genes known to regulate immune response included downstream targets of *HIF1A* that influenced our GSEA results, such as *EGFR*, which regulates immune response through *EGR1* (53, 54), and *CXXC5* (55). It has also been suggested that hypoxia triggers neutrophil degranulation (one of the differentially targeted immune response pathways), which in turn leads to increased necrosis and poorer survival outcomes in GBM (56). These hypoxia pathways likely regulate immune response in tandem with immune regulatory processes at baseline, which are known to exhibit sex differences (18). To improve efficacy of chemotherapy in females with GBM by modulating the effects of *HIF1A* on these downstream processes, therapies that target HIF in RCC, such as those discussed by Golijanin *et al*. (57), should be explored as possible protocols for sex-specific GBM treatment.

The GBM-specific targeting of the spliceosome in males, but not in females, that we found is consistent with previous studies that have found that different TFs target the spliceosome in the healthy brain tissue of males and females, which likely contributes to the sex-specific differential targeting of these pathways in GBM (18). Alternative splicing is also known to adversely affect treatment response independent of hypoxia by altering key molecular functions that include transcription factor activity, in turn, by increasing angiogenesis and facilitating cell migration (58-60).

We note that this work has some limitations. First, while we used PANDA to estimate GRNs, the method does not indicate whether a regulatory relationship is activating or inhibitory, and this can complicate interpretation. Furthermore, because the data were collected at a single time point in each individual, causal relationships must be estimated using prior knowledge rather than inferred from causal models. The GBM and LGG categories were assigned in TCGA according to the 2016 WHO classification of gliomas (61), not the 2021 update (2); this may result in some gliomas being misclassified, although the differences between these extremes are likely to be relatively minor. Further, it is unclear whether the differential targeting of immune processes such as neutrophil degranulation is due to differing immune cell activity by sex rather than differential abundance of these immune cells overall; the second scenario may occur due to alterations to the blood-brain barrier that are linked to angiogenesis or immune processes regulated by *HIF1A* (62, 63). Finally, representation in TCGA is an issue. TCGA samples are highly skewed towards people of European descent, and we also observe data imbalance (64), with the GBM-F group having fewer samples than the GBM-M, LGG-M, or LGG-F groups. This small sample size complicates generalization to the whole GBM-F population. However, because our analyses are based on pairwise correlations, we do not expect the sample size to affect statistical power.

### Perspectives and significance

The analyses presented here represent an advance in our understanding of GBM-specific mechanisms in males and females. The sex-specific differential regulation we uncovered sheds light on the importance of hypoxia and the differential regulation of downstream pathways in GBM-specific treatment resistance in females, and suggests exploring *HIF1A* targeting as a potential avenue for GBM treatment in females. Our analysis also indicates there may be alternative mechanisms causing treatment resistance in males, a result that could be further explored to develop therapeutic strategies for managing the disease in males.

## Conclusions

We used a set of complementary methods to analyze GRNs in LGG and GBM with the goal of understanding sex-biases in clinical outcomes, including response to therapy. This included a combination of pathway enrichment analysis, GRN differential targeting analysis, and GRN co-regulation analysis. We used the differences between LGG and GBM as a proxy for treatment resistance in GBM based on the overall better response and longer survival of individuals with LGG independent of sex. We found that regulation of hypoxia pathways, and co-regulation of hypoxia pathways with carbohydrate metabolism, extracellular matrix, and immune pathways, was increased in females with GBM both in comparison to males with GBM and females with LGG. In males with GBM, we found increased co-regulation of carbohydrate metabolism, extracellular matrix, and immune pathways with the formation and disassembly of the mRNA spliceosome and increased regulation of mRNA spliceosome and androgen receptor pathways in comparison to both females with GBM and males with LGG.

## Data Availability

Gene expression data for GBM and LGG are accessible through Zenodo (https://zenodo.org/records/18226833), the protein-protein interaction data used for GRN inference are available from the GRAND database (https://granddb.s3.amazonaws.com/tissues/ppi/tissues_ppi.txt), the curated gene set file used for pathway analysis is available from MSigDB (https://www.gsea-msigdb.org/gsea/msigdb/collections.jsp), and the basic gene annotation file used to subset protein coding genes is available from GENCODE (https://www.gencodegenes.org/human/). Male-specific and female-specific motif binding data, clinical data for GBM and LGG, and code to replicate analysis are available from GitHub (https://github.com/QuackenbushLab/Adebari_Glioma_scripts).

https://granddb.s3.amazonaws.com/tissues/ppi/tissues_ppi.txt

https://www.gsea-msigdb.org/gsea/msigdb/collections.jsp

https://www.gencodegenes.org/human/

https://github.com/QuackenbushLab/Adebari_Glioma_scripts

## Declarations

### Ethics approval and consent to participate

Data were accessed through a public portal made available as part of the Cancer Genome Atlas Project (TCGA) and are subject to TCGA ethical guidelines and policies.

### Consent for publication

Not applicable

### Authors’ contributions

TE and JQ conceptualized the project. TA and TE performed the analysis. VF assisted TA with use of NetZooPy, MBG assisted TA with use of GRAND, and DD assisted TA with use of WebMeV. CLR provided sex-specific motif priors for PANDA analysis. LH developed a custom function for MONSTER for use in this analysis. TA, TE, and JQ wrote the manuscript. KHS, DLD, and CLR provided feedback on project scope, analytical techniques, and presentation of results.

## Acknowledgements

We thank Roaa Ben Aiad for feedback regarding the figures.

## Supporting information

**S1 Table**. PANDA results for LGG (male).

**S2 Table**. PANDA results for LGG (female).

**S3 Table:** PANDA results for GBM (male).

**S4 Table:** PANDA results for GBM (female).

**S5 Table**. Pathway analysis results for LGG male vs. LGG female.

**S6 Table**. Pathway analysis results for LGG female vs. LGG male.

**S7 Table**. Pathway analysis results for GBM male vs. GBM female.

**S8 Table**. Pathway analysis results for GBM female vs. GBM male.

**S9 Table**. Pathway analysis results for LGG male vs. GBM male.

**S10 Table**. Pathway analysis results for GBM male vs. LGG male.

**S11 Table**. Pathway analysis results for LGG female vs. GBM female.

**S12 Table**. Pathway analysis results for GBM female vs. LGG female.

**S13 Table**. MONSTER results for LGG female vs. GBM female.

**S14 Table**. MONSTER results for LGG male vs. GBM male.

**S15 Table**. Annotated significant TFs in LGG to GBM rewiring.

**S16 Table**. BLOBFISH network specific to GBM female, consolidated by pathway.

**S17 Table**. BLOBFISH network specific to GBM male, consolidated by pathway.

